# OGSCalc: Mathematical formulae and web-based application to incorporate rotational discrepancies into translational discrepancies for assessment of accuracy in orthognathic surgery

**DOI:** 10.64898/2026.04.03.26350094

**Authors:** Jonas Hue, Joella Yeo, Leonardo Saigo

## Abstract

**Objectives:** Accurate assessment of orthognathic surgical accuracy is essential in the evaluation of operative techniques. Surgical accuracy is often reported as rotational and translational deviations from planned positions. This results in 6 separate values, translation in three planes, anterior-posterior (AP), superior-inferior (SI) and medial-lateral (ML) and rotations—pitch, roll and yaw. However, rotations will influence 3-dimensional positions and translational discrepancies.

**Methods:** We have derived a mathematical formula using Euclidean geometry and quadratic functions that quantifies the impact of rotations on translational discrepancies. This allows for the calculation of a total discrepancy value that incorporates the three translations and rotations. Furthermore, we developed an interactive web-based application using the open-source shiny R package.

**Results:** We have successfully reduced equations from Euclidean geometry into a quadratic form. The equation is as follows, [4(sinθ/2)2-2]x^2^ + [8d(sinθ/2)2-2d]x + 4d2(sinθ/2)2 = 0, where θ represents the rotational discrepancy in radians and d represents the translation discrepancy. This allows us to solve for the correction needed to be made to translational discrepancies to account for the influence of rotational discrepancies.

We successfully developed a web application with a user-friendly graphical user interface. Clinicians upload their own data in the excel (.xlsx) file format and the application automatically performs the necessary calculations over many patients, returning a downloadable table of results.

**Conclusion:** We present a mathematical formula incorporated into a web-application to combine translational and rotational discrepancies for deeper insight when evaluating orthognathic surgical accuracy.

**Clinical Relevance:** This allows surgeons to account for rotational influence on 3-dimensional translational discrepancies.

## Introduction

The assessment of orthognathic surgical accuracy is essential in evaluating the success of the surgery and comparing different surgical techniques. This can be performed by comparing the actualised changes after orthognathic surgery to the virtual surgical plan. In the past, post-surgical assessments were often performed with 2D lateral radiographs^1^. However, advances in 3D image analysis have allowed surgeons to accurately quantify their surgical accuracy via measurements of translational and rotational discrepancies from the planned positions. These values are often reported as translational discrepancies in three axes, anterior-posterior (AP), medial-lateral (ML) and superior-inferior (SI). A further three values are reported for rotational discrepancies about three axes, pitch, roll and yaw^2^. Various software programmes are available for semi-automated registration of planned and post-operative imaging. These programmes can be used to calculate translational and rotational discrepancies. One such programme that has been gaining popularity is the OrthoGnathicAnalyzer^3,4^.

Unfortunately, these 6 values (three translation and three rotations) are often reported as individual and separate numbers. This makes comparison between surgical techniques very difficult unless all six values are in agreement. Furthermore, we understand the translation and rotation are inherently linked. For example, an AP discrepancy is reduced by pitch and yaw rotations as rotations about these axes bring a reference point closer to the reference plane. Each translational discrepancy is influenced by rotation about two axes. ML discrepancy is influenced by yaw and roll while SI discrepancy is affected by pitch and roll. Despite their inter-linked relationships, these discrepancies are often analysed separately.

Therefore, in this work, we present a series of mathematical formulae to derive a single, unified discrepancy value based on the three translational and three rotational discrepancies. This calculation corrects for the influence of rotation on translational discrepancies.

The existing literature and some of our previous work have shown the utility of simple scripts to automate repetitive tasks which can be applied to a wide range of scenarios from basic science research^5,6^ to clinical studies^7^. These tools can save researchers and clinicians time and facilitate efficient analysis of data. However, these tools should be user-friendly and ideally built on open-source platforms with code made publicly available for academic scrutiny and future adaptations. Therefore, as the formulae are not easily calculated by hand, we have further developed a web-application which allows the clinician to upload their own data and the application performs the calculations automatically and efficiently.

## Methods

### Calculations

In order to incorporate the influence of rotational discrepancies into translational discrepancies, we have utilized a combination of formulae for chord lengths and Pythagoras theorem, to reduce the equations to a quadratic form. The quadratic formula is then applied to solve for the correction required for each translational discrepancy in the presence of rotational discrepancies. For simplicity, Figure 1 depicts how rotations influence the calculated translational discrepancies in one dimension. Taking the AP dimension as an example, a pitch angle of θ results in a reduction in the AP discrepancy (*d*) by a distance of x. Furthermore, AP discrepancy is also affected similarly by yaw. Therefore, the AP discrepancy needs to be corrected for rotations of pitch and yaw. ML discrepancy needs to be corrected for yaw and roll while SI must be corrected for pitch and roll.

**Figure 1.**
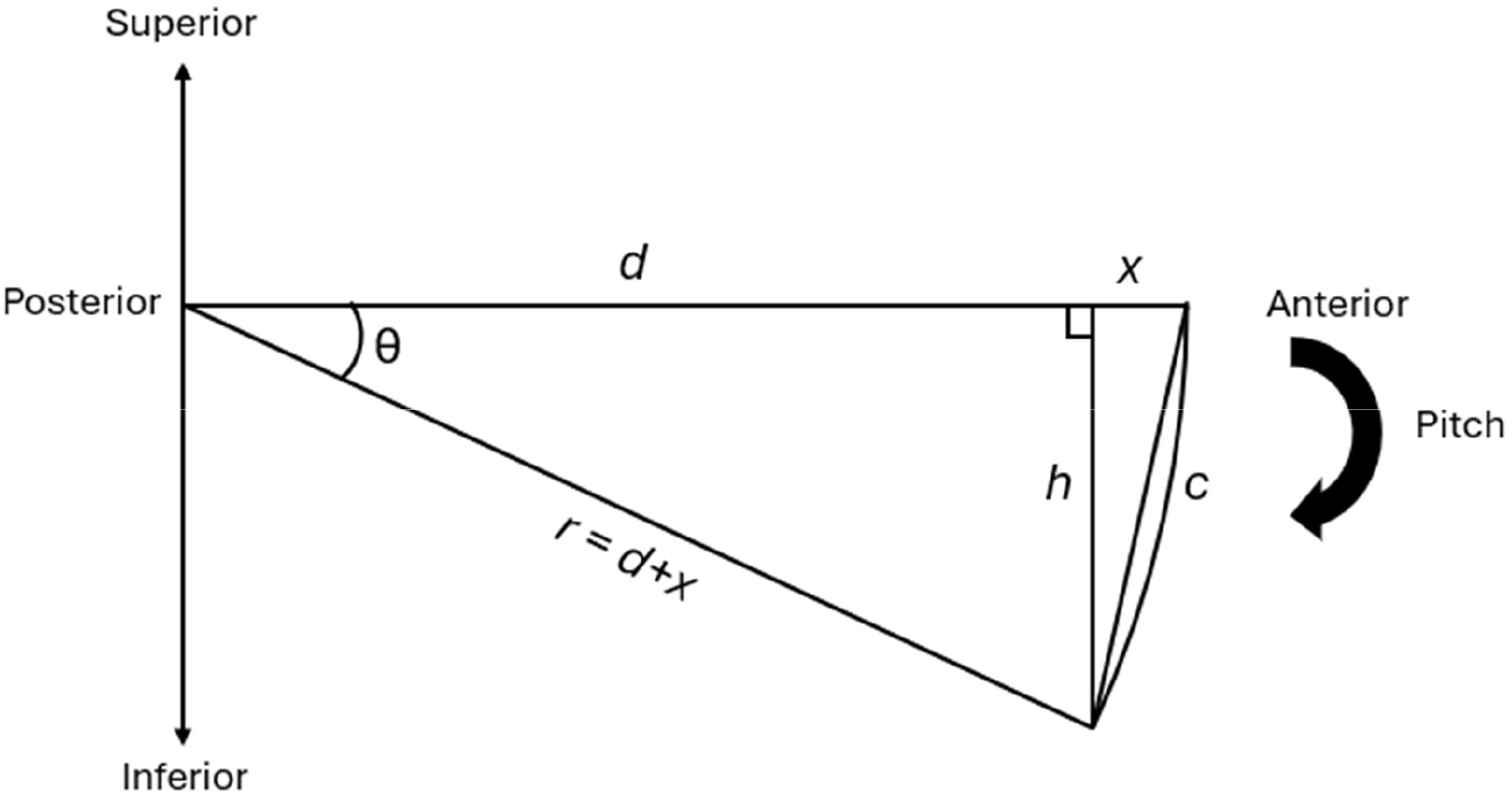
Schematic depicting the influence of rotations (θ) on translational discrepancy (*d*) using the Anterior-Posterior dimension and pitch as an example. Chord length (*c*) forms the hypothenuse of one right-angled triangle with base, *x*, and height, *h*, where *x* is the correction to be calculated to obtain the true translational discrepancy.

Figure 1 illustrates the geometric diagram and various known and unknown distances for the calculation. First, we start with the formula for chord lengths as follows:

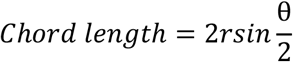

Where *c* represents the chord length, *r* is the radius of the circle, represented by the true discrepancy (*d* + x) and θ is the angle of the rotational discrepancy about 1 axis. There are two right angled triangles each with height *h*.

Therefore, using Pythagorean theorem, we can write two equations as follows:

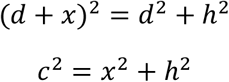

Inputting the formula for chord length into the second equation, we get the following:

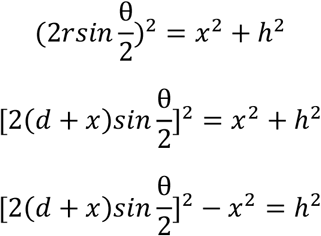

This can then be combined with the first equation with the common unknown, *h*^2^, to obtain the following:

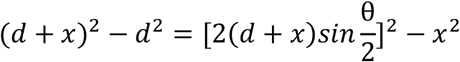

This equation can then be reduced to a quadratic form in the following steps:

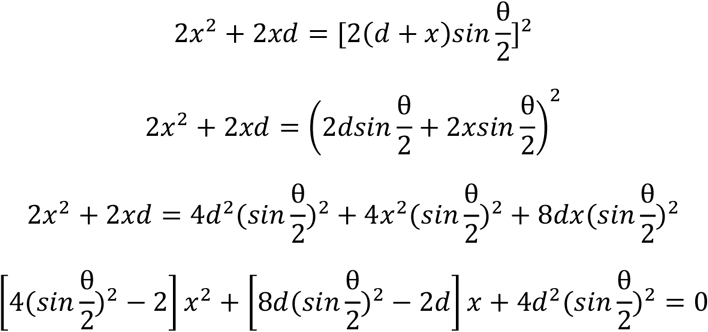

Using the quadratic formula, we are able to solve for the discrepancy correction, *x*, with known values for translational discrepancy, *d*, and rotational discrepancy, θ.

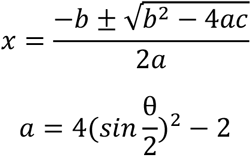

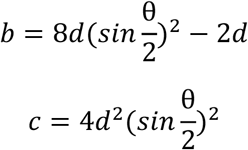

This process is repeated twice for each of the rotational axes that influence the translational discrepancy in each dimension. With the corrected, translational discrepancies, we can calculate a true total discrepancy with the formula for Euclidean distance below. This value represents inputs from the 6 original data points—3 rotations and 3 translations.

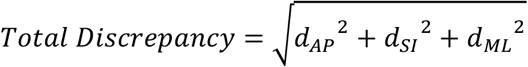

### Web Application

A web-based application was built in the R programming language (version 4.4.2) using the shiny^8^ package. The application will be deployed at https://jonashue.shinyapps.io/OGSCalc/ and the source code made publicly available on GitHub at https://github.com/jonashue1/OGSCalc.

The application was created to be simple and easy to use. The application allows users to upload their own cohort of data as an Excel Open XML Format Spreadsheet (.xlsx) file (Figure 2A). The data should be organised with 7 columns in the following order—case number, AP, SI, ML, pitch, roll and yaw discrepancies (Table 1). The user then has to specify the number of patients (Figure 2B) in the cohort and how many decimal places (Figure 2C) they would like the calculator to return the output with. Finally, users can download the calculated values in a comma-separated values (.csv) file (Figure 2D).

**Table 1.** Table of example data illustrating the format for users to upload their own .xlsx files. The data should have seven columns—case number, AP, SI, ML discrepancies and pitch, roll and yaw discrepancies.

| CaseID | AP | SI | ML | Pitch | Roll | Yaw |
| --- | --- | --- | --- | --- | --- | --- |
| A1 | 1.6987 | 1.5449 | 2.1495 | 1.9362 | 1.6492 | 2.3976 |
| A2 | 2.0789 | 1.5481 | 3.1112 | 3.2302 | 2.4016 | 1.2353 |
| A3 | 1.9526 | 1.3992 | 2.7096 | 1.9034 | 0.8116 | 2.9610 |
| A4 | 2.5321 | 1.8699 | 3.1187 | 1.9222 | 2.0114 | 4.5077 |
| A5 | 2.0702 | 1.5617 | 1.8485 | 1.5170 | 1.4389 | 0.7553 |
| B1 | 2.1912 | 1.4853 | 2.1492 | 1.8447 | 2.3611 | 3.6824 |
| B2 | 1.6509 | 1.3056 | 1.9238 | 2.1280 | 1.5183 | 1.5411 |
| B3 | 2.4287 | 1.7554 | 2.6848 | 2.2921 | 2.3595 | 4.4424 |
| B4 | 1.5048 | 1.0431 | 1.5738 | 2.7458 | 1.7219 | 4.0432 |
| B5 | 1.7841 | 2.6551 | 2.6977 | 2.6791 | 0.7607 | 2.0450 |

**Figure 2.**
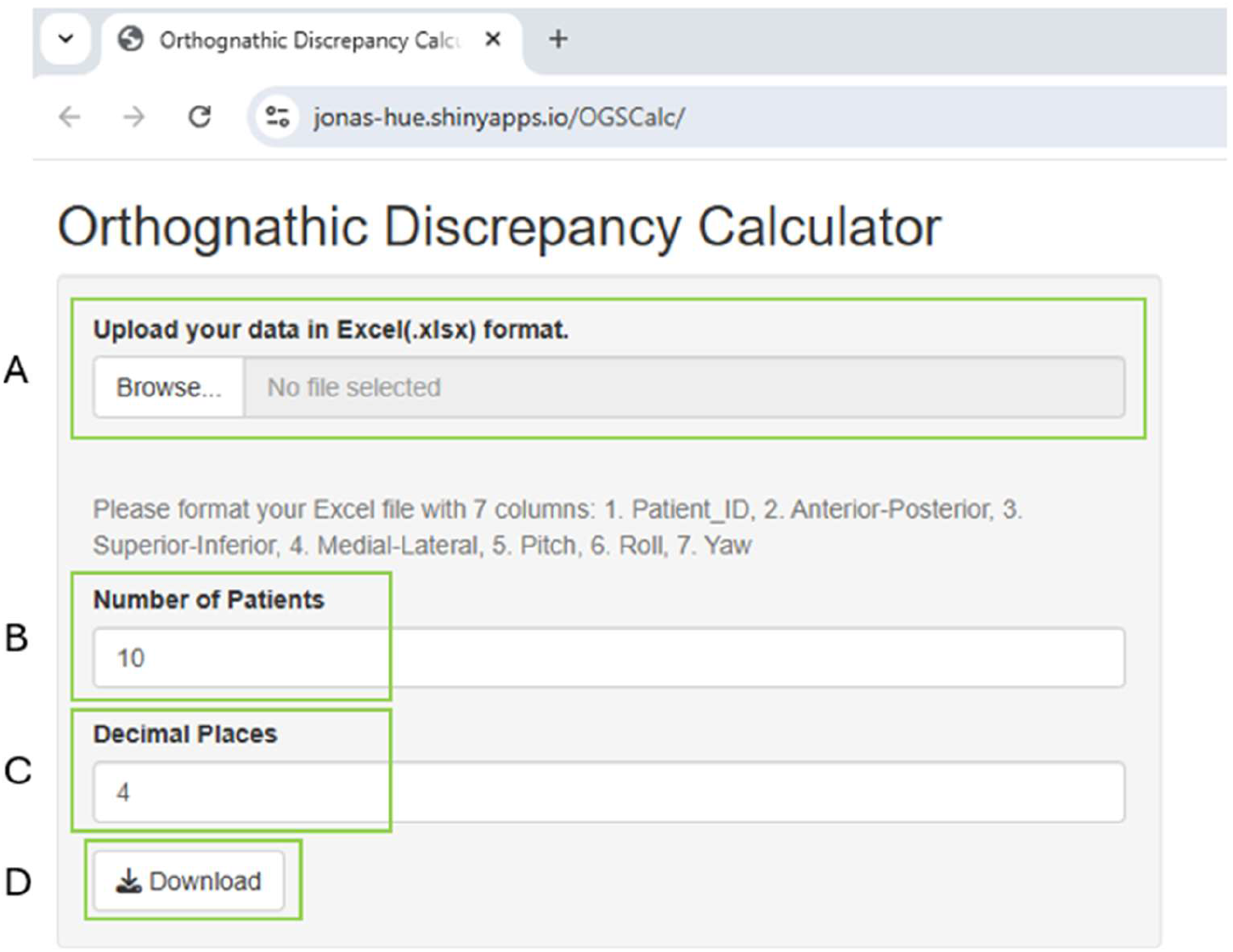
User interface of the OGSCalc web application. (A) File upload button for users to upload their own .xlsx files. (B) Specify the number of patients in the cohort for analysis. (C) Specify the number of decimal places for the calculated total discrepancy. (D) Download button for the calculations in .csv format.

The application is structed into four main components. The required R packages, the various functions to perform the calculations, the user interface and server functions. First, the required libraries are loaded which includes shiny, readxl, tidyverse and DT. The readxl package facilitates the uploading of .xlsx files from the user. The DT package creates a HTML widget to allow data objects in R to be displayed as tables on HTML pages.

Subsequently, the various formulae and equations as described above are then translated into various functions. This allows us the application to call these functions each time a calculation is required.

The user interface component creates the visual elements of the application that the user interacts with (Figure 2). This includes a file upload control which allows users to upload their own data. Two numeric input boxes are also added to allow users to specify the number of patients and decimal places for the calculated output. Finally, a download button is added to allow users to download the OGSCalc’s output.

The server function runs the required processing steps for the web-application to return the discrepancy calculations. Reactive expressions are written to pull the data uploaded by the user. The uncorrected translational and rotational discrepancies are used to calculate the total, corrected discrepancy using the various functions written previously. A set of tabs are created to allow users to toggle between showing the raw, uploaded dataset and the table of calculations (Figure 3).

**Figure 3.**
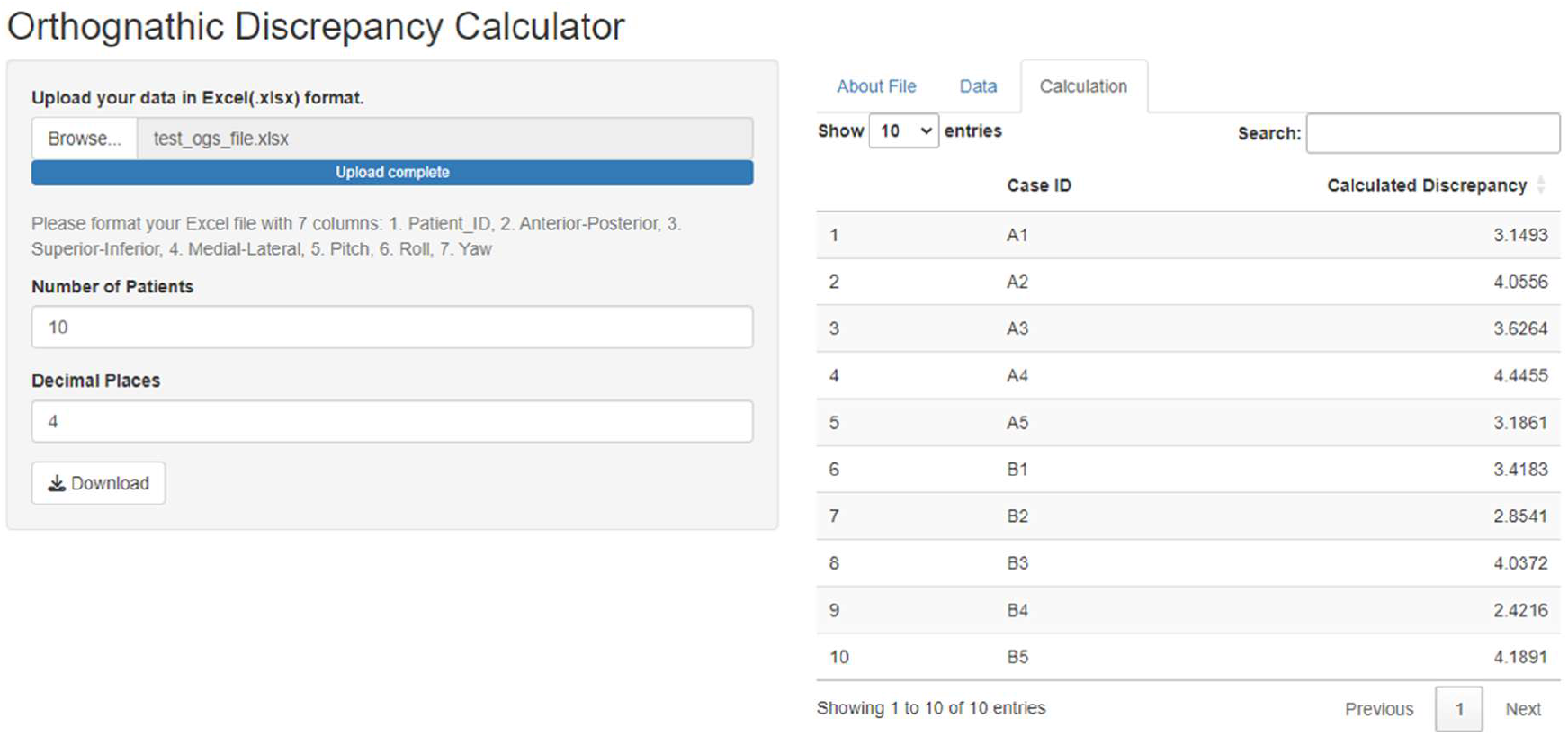
OGSCalc web application in function, showing the calculations returned after the upload of sample data. The application has different tabs allowing users to toggle between panels to explore their dataset and calculated outputs.

## Results

The web-based Shiny application is successfully deployed at the following uniform resource locator (URL): https://jonas-hue.shinyapps.io/OGSCalc/. The user interface and different aspects of the OGSCalc application as described in the methods are shown in Figure 2.

To showcase the application in function, we generated some normally distributed example data in R using the rnorm function. We created a cohort of 10 example patients with mean translational discrepancies of 1.5-2.0mm and standard deviation (SD) of ±0.5-0.8mm and rotational discrepancies of 1.7-2.8° with SD of ±0.5-0.9°. The data and appropriate format for upload is shown in Table 1. Users may take reference from this table to format their own cohort of patients before using the application.

The data in Table 1 was uploaded to the OGSCalc application. Figure 3 shows the working application. The data was successfully uploaded as an .xlsx file containing a total of 10 patients. The 6 discrepancy values for translation and rotation are used to calculate the total, corrected discrepancy as shown in the “Calculation” tab in the application (Figure 3). Within this panel, users can search through their data and filter or order the data as necessary.

Using the data shown in Table 1, we calculated the Pearson’s correlation between each of the uncorrected translational discrepancies with output from OGSCalc. AP discrepancy had a correlation of 0.741, 95% CI [0.209, 0.935]. SI discrepancy had a correlation of 0.732, 95% CI [0.191, 0.932]. ML discrepancy had a correlation of 0.938, 95% CI [0.752, 0.985]. This shows the wide variation between correlation of translational discrepancies in each dimension, supporting the reporting of a corrected total discrepancy value.

## Discussion

In this work, we have derived a formula to account for the influence of rotations on translational discrepancies in orthognathic surgery. To the best of our knowledge, no such formulae have been previously described or employed in the literature. While the formula is relatively simple, it provides clinicians a method to accurately calculate discrepancies from the surgical plan and assess accuracy in orthognathic surgery. Prior to this work, while we know rotations will influence translations and 3D positioning of reference points, there is no method to quantify their impact on translational discrepancies. Furthermore, given that rotational discrepancies vary from patient to patient, it becomes impossible to directly compare translational discrepancies without properly accounting for these rotations.

Over the years, new techniques have been introduced in orthognathic surgery from novel osteotomies^9^ to computer-assisted surgery^10,11^ to mixed reality^12^ and navigation^13-15^ guided surgery. This emphasizes the need for reliable and standardised methods to compare these to conventional surgical methodology^16^. Previous studies often report translations in 3 dimensions and rotations about 3 axes as 6 separate values when evaluating accuracy^17-19^. However, unless all 6 values are in agreement and superior in one technique compared to controls, it becomes difficult to draw conclusions about the new techniques. As we have seen in the literature, such clear-cut findings are rarely the case^2,13,20^. We often see certain techniques having significantly different discrepancy in just one or two dimensions or axes. For example, in the study by Schrader et al. (2025), surgical navigation significantly improved orthognathic surgical accuracy, but only in the ML and SI dimensions and yaw^13^. Since yaw also affects ML translation, the control group with a higher yaw discrepancy would also have a stronger rotational impact on ML discrepancy. Hence, correcting for rotational discrepancies becomes especially crucial where there are significant differences in certain rotational discrepancies as the translations will be differentially influenced by the rotations. Therefore, the formulae presented here allow us to make corrections for these different rotations which facilitates more accurate and direct comparison of translational discrepancies.

Our simulated data has also shown how translational discrepancies in each dimension can vary in correlation to the overall 3D distance. While they are inter-linked by the distance formula, there may be substantial differences between their correlations to overall discrepancy. Therefore, this emphasizes the importance of continued reporting of discrepancies in each of the 3 dimensions and 3 rotations in addition to the output from OGSCalc when reporting surgical accuracy. The aim of our tool is to provide an adjunctive metric to better assess orthognathic surgical accuracy rather than to replace existing methods.

It is also important to note that as novel surgical techniques incrementally improve accuracy, rotational inaccuracies will also reduce over time. As the rotational discrepancies get smaller and smaller, the corrections made will also gradually approach zero. We hope that in the future, OGSCalc will gradually become obsolete as our surgical techniques improve. Nonetheless, the formulae and web application offer clinicians a method to quantify the influence of rotations on translations and check if a correction is required prior to data analysis.

Finally, the application we have created is built on open-source platforms with the code made publicly available. We believe in the importance of democratization of data where clinicians and researchers can adopt and adapt our work for their own purposes. Our code will remain public for transparency and academic scrutiny. We hope that researchers will find this useful and contribute to improvements in our simple tool. The tool runs well on the shinyapps.io platform and returns calculations almost immediately. We have employed the OGSCalc application to our own cohort of patients in a study comparing navigation to conventional occlusal wafer techniques (unpublished data, manuscript submitted for publication). We hope that other clinicians will also find utility in our web application.

## Conclusion

In conclusion, we have derived a series of mathematical formulae which allows us to account for the influence of rotations on translational discrepancies in orthognathic surgery. We have also developed a web-based shiny application to create a user-friendly interface for clinicians to perform efficient and automated calculations on their own datasets.

## Data Availability

All data produced in the present study are available upon reasonable request to the authors.

https://jonas-hue.shinyapps.io/OGSCalc/

https://github.com/jonashue1/OGSCalc

